# A Vaccination Simulator for COVID-19: Effective and Sterilizing Immunization Cases

**DOI:** 10.1101/2021.03.28.21254468

**Authors:** Aknur Karabay, Askat Kuzdeuov, Shyryn Ospanova, Michael Lewis, Huseyin Atakan Varol

## Abstract

Accurate modeling provides a means by which a complex problem can be examined for informed decision-making. We present a particle-based SEIR epidemic simulator as a tool to assess the impact of vaccination strategies on viral propagation and to model both sterilizing and effective immunization outcomes. The simulator includes modules to support contact tracing of the interactions amongst individuals as well as epidemiological testing of the general population. The simulator particles are distinguished by age, thus enabling a more accurate representation of the rates of infection and mortality in accordance with differential demographic susceptibilities and medical outcomes. Moreover, thanks to the age differentiation of particles, the vaccination can be simulated based on the age group descending order or randomly across all age groups. The simulator can be calibrated by region of interest and variable vaccination strategies (i.e. random or prioritized by age) so as to enable locality-sensitive virus mitigation policy measures and resource allocation. The results described, based on the experience of the province of Lecco, Italy, indicate that the tool can be used to evaluate vaccination strategies in a way that incorporates local circumstances of viral propagation and demographic susceptibilities. Further, the simulator accounts for modeling the distinction between sterilizing immunization, in which immunized people are no longer contagious, and that of effective immunization, in which symptoms and mortality outcomes are diminished but individuals can still transmit the virus. The sterilizing-age-based vaccination scenario results in the least number of deaths compared to other scenarios. Furthermore, the results show that the vaccination of the most vulnerable portion of the population should be prioritized for the effective immunization case. As the vaccination rate increases, the mortality gap between the scenarios shrinks.

## I. Introduction

The scope and scale of the COVID-19 epidemic are unprecedented in modern times. As a novel coronavirus with a high rate of propagation, its emergence caught the world unprepared, with little or no natural immunity amongst the population, and no existing effective vaccine. The impact has been staggering, in both social and economic terms. The global health toll to date (March 2021) is in excess of 120 million confirmed cases and more than 2.7 million deaths [1], exhausting healthcare systems and presenting significant concern about long-term health impact even for those who experienced mild cases. Initial attempts to control the spread have damaged national economies and disrupted global trade, thus exposing the vulnerabilities of existing systems, and serving to erode public confidence in and adherence to government efforts to bring the situation under control.

The main first-line methods used by governments to suppress the spread and mitigate the impact consisted primarily of non-pharmaceutical interventions (NPIs) such as social distancing, the use of masks, the restriction (or even closure) of non-essential businesses, the widespread adoption of remote-work policies, an abrupt transition to online education, and on occasion, the imposition of more severe physical quarantine restrictions.

However, the widespread and uneven implementation of NPIs caused significant social disruption and negative economic outcomes. At the moment, it became imperative to consider the full range of intervention strategies, in order to identify the optimal combination of NPIs to maximize benefits while minimizing negative repercussions. To these ends, a number of epidemic simulators were developed (compartmental, agent-based, and particle-based) to assess the effectiveness of available measures, and to consider the trade-offs associated with the governmental policies and resulting economic and social impact [2]–[10].

The recent emergence of vaccines is a welcome development, as it introduced more effective pharmaceutical responses to the policy mix. The vaccine-based approach is expected to become the dominant strategy, gradually diminishing the reliance on the NPIs and their associated disruptions. Vaccines such as Pfizer-BioNTech COVID-19, Moderna COVID-19, Sputnik V completed their clinical studies and received regulatory approval for public use in many countries [11]–[13]. Initial results indicate high efficacy rates for these vaccines, usually above 90%, after the two-shot individual regimen is administered.

The effective large-scale deployment of the vaccines can significantly increase the percentage of the population exhibiting a level of immunity to the virus. Combined with those previously infected, and thus possessing a degree of natural immunity, it now becomes feasible for communities to achieve levels of immunity sufficient to be described as “herd immunity”, which is considered the surest way to suppress the epidemic, ongoing, and to protect the most vulnerable groups of the society [14], [15]. Herd immunity is achieved when the pool of those remaining vulnerable to infection is sufficiently small as to reduce the propagation rate (R-nought) well below the threshold value of 1, at which point the virus gradually diminishes naturally, and thereby reduce the risk of infection for those who cannot be vaccinated (e.g., children and people with certain medical conditions) [14]. Although herd immunity can be attained through both natural exposure to the infection and vaccination of the individuals, the latter is preferred since it results in fewer deaths and minimizes the pressure on the healthcare system [15].

The vaccines for COVID-19 were developed at an accelerated pace, much faster than usual, in a race against time. The type and duration of immunization that they can provide against SARS-CoV-2 are still unknown [16]. Vaccine developers aspire to achieve the outcome of “sterilizing immunization”, which stops the reproduction of the virus in the body such that the person can no longer become contagious, though sterilizing immunity has rarely been achieved [7], [16]. In most cases, a vaccine primarily prevents the development of severe symptoms upon exposure, while the patient may remain contagious; we describe this as “effective immunity”. To emphasize the point, attaining effective immunity does not imply that further transmission of the disease by the vaccinated individual is prevented [17]. Thus, the type of immunization achieved by vaccination needs to be evaluated, as well as the transmission rate, to assess the course of the different vaccination strategies, which requires monitoring and observation over time. Furthermore, whether the vaccination reduces the severe cases or transmission rates of the infection is critical in determining the optimal vaccination strategy [18]. Even with a high number of daily vaccines per thousand people and efficient distribution it will take considerable time to achieve herd immunity levels, likely extending into 2022 and beyond. This puts additional concerns on the new variants arising with time for which the existing vaccines might not be as effective as for the existing variants [19]. Thus, it is imperative to devise vaccination strategies that optimize the desired outcomes for the short- and middle-term and do so in a manner that is responsive to local conditions. Operational challenges such as production bottlenecks, logistics, storage requirements as well as accurate categorization of vaccination priority groups and economic issues are critical. The situation is compounded by the fact that many of the most prominent vaccines require a second dose to achieve the desired level of immunity, a requirement that adds additional burdens of record-keeping and follow-up to ensure timely delivery of the second dose [20].

Beyond the medical and logistical challenges, there is the issue of public sentiment regarding vaccines. Vaccine misinformation has spread widely in an online environment that is largely unregulated. Thus, informing and educating the public is crucial to achieving the desired high rates of vaccination among the population. Several surveys have been conducted to investigate the public perception of vaccination. For example, vaccine hesitancy was reported as 64%, 67%, and 75% in the United Kingdom [21], the United States [22], and Israel [23], respectively. However, the vaccine acceptance rate is much higher among high-risk groups such as the elderly and healthcare workers. Moreover, compulsory vaccination might reduce the vaccine uptake, while other factors such as vaccination cost and vaccine efficacy might considerably influence the vaccine uptake rates [18]. In this context, epidemic modeling is essential to generate accurate estimates and develop optimized strategies taking into account the aforementioned conditions. In our earlier work, we developed a particle-based COVID-19 susceptible-exposed-infectious-recovered (SEIR) simulator with modules for contact tracing and epidemiological testing [4]. The simulator was successfully used to model the epidemic spread under different rates of testing and contact tracing of infected people at the individual level. In this work, we extend this simulator by adding a vaccination module with modes for both effective and sterilizing immunization. The tool can be used to simulate age-based vaccination strategies as well as randomized vaccination of the population. This enables us to observe the relation between vaccination rates and population prioritization, and monitor the progression of overall vaccination strategies.

The rest of the paper is structured as follows: Section II presents the review of the works on vaccination simulators and highlights the comparative novelty of our work. Section III gives an overview of the particle-based model, the methodology of the particle-based SEIR simulator, and the vaccination module. Simulations for a validation scenario and different vaccination strategies are provided in Section IV. The results are evaluated and discussed in Section V and Section VI concludes the paper with directions for future works.

## II. Related Work

The COVID-19 vaccine rollout has begun, representing a historic milestone but simultaneously generating considerable debate regarding strategy and how best to achieve the desired goals. The circumstances demonstrate the acute need for an effective COVID-19 vaccination simulator as a tool to model varying scenarios, and thereby contribute to the development, deployment, and ongoing review of vaccination rollout strategies. The need for ongoing review is emphasized, due to the uncertainties in the transmission parameters, longevity of the immunity, and immunity type after vaccination. Given this motivation and purpose, researchers are actively working on the development of various simulators that can be used to model vaccination strategies for COVID-19 [5]–[10].

Agent-based COVID-19 simulators have been proposed in [5], [9], [24]. [5] uses SIR model and divides the population based on age groups. Each agent is assigned an arbitrary schedule covering different locations such as school, home, and work that have different transmission intensity. The model is initialized with different ratios of recovered to susceptible individuals, where the recovered portion represents both recovered and vaccinated populations. Based on different initial vaccinated portions of the population, the future course of the epidemic can be predicted, along with an analysis of the vaccination coverage required to achieve herd immunity.

In [22], the SEIR model with the vaccination module was presented where the population was divided into three categories representing different health states associated with the socio-economic classes of society. The authors considered two vaccination strategies: random vaccination of all three groups and prioritization of the second and third categories that represent mild and severe medical conditions. Patel et al. created an agent-based SEIR model where agents were differentiated by age which was linked to the mortality and hospitalization rates, respectively [24]. The authors introduced different vaccination coverage with various efficiencies. The vaccinated agents were moved to the recovered state which modeled the sterilizing immunization case. COVID-19 simulators based on compartmental models were also developed [20, 23, 25]. Researchers created a SimCOVID tool that used compartmental SIR, SEIR, and SEIRD compartmental models [23]. The authors used the sigmoid function to smooth the output of a fine-tuned model that was based on the daily registered COVID-19 cases.

MacIntyre et al. [6] presented a deterministic compartmental SEIR model that also divides the population based on age in order to model infection spread and vaccination strategies; this model assumed two-stage vaccination. The age range of the population was considered to be from 0 to 80 years old, and this was split into 16 five-year groups. Age groups were used to account for varying rates of transmission across the groups. Different vaccine efficacy rates were simulated to analyze the coverage required to achieve herd immunity. In this model, only susceptible and pre-symptomatic individuals were vaccinated.

Another research group developed an SIR compartmental model for simulating vaccination strategies. This simulator assumed a sterilizing immunity after one-dose vaccination, which implies no virus reproduction and transmission [25]. It is not clear yet whether sterilizing immunity will be obtained, as mentioned earlier [7]. The model incorporated both dead and recovered individuals in the removed state. The authors performed simulations for different rates of vaccine efficacy with a constant vaccination rate.

Our work is unique in that it enables simulation of both effective and sterilizing immunity obtained after vaccination, using either a one or two-shot delivery. In addition, our particle-based SEIR simulator can model the effects of multiple strategies (contact tracing, testing, and vaccination) concurrently. The target population can be restricted by age groups to enable prioritization based on the considered scenario. Each vaccination module can be run in two modes: age-based and randomized vaccination of the population. Furthermore, the tool considers vaccination of the recovered, exposed and infected (representing asymptomatic cases) along with the susceptible which more accurately imitates the real scenario. Therefore, by varying the aforementioned parameters the relation between target population, vaccination immunization type and vaccination rate can be analyzed. The strategies can be examined on a more granular basis, by region, considering the three main aspects of decreasing total mortality, minimizing pressure on the healthcare system, and resources available for vaccination [8]. These three criteria can be evaluated in our simulator by selecting the strategy, vaccination mode, and daily vaccination rate.

## III. Methodology

This work is an extension of prior work on particle-based simulator with contact tracing and testing modules. Thus, the methodology will cover the particle model, with a description of the SEIR simulator, and the vaccination module. The reader is referred to [4] for detailed descriptions of the contact tracing and random testing modules.

### A. Particle Model

In our simulator, each individual is modeled as a particle *p* which is represented as a vector:

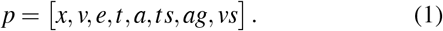

The definition of each element in this vector is provided in Table I.

**TABLE I:**
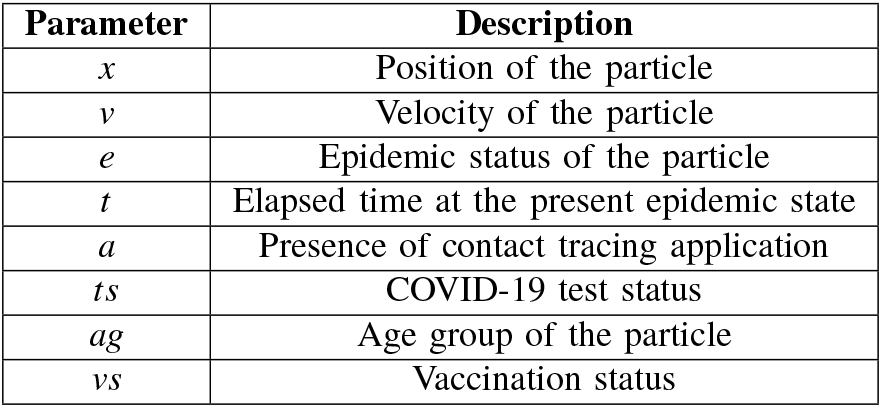
Particle model parameters and their definitions.

| Parameter | Description |
| --- | --- |
| $x$ | Position of the particle |
| $v$ | Velocity of the particle |
| $e$ | Epidemic status of the particle |
| $t$ | Elapsed time at the present epidemic state |
| $a$ | Presence of contact tracing application |
| $ts$ | COVID-19 test status |
| $ag$ | Age group of the particle |
| $vs$ | Vaccination status |

The motion and interaction of the particle are modelled on a two-dimensional map. At the start, the velocity and position values of all *n* particles are initialized based on the constraints −*v*_*max*_ ≤ *v*_*i j*_ ≤ *v*_*max*_ and −1 ≤ *x*_*i j*_ ≤ 1, where *i* = 1, …, *n* is particle number and *j* = 1, 2 refers to the two dimensions of the map. The position and velocity of the particles are stored in the matrices *X* ∈ℝ^*nx*2^ and *V* ∈ℝ^*nx*2^, respectively. At every iteration κ (1 ≤ κ ≤ *T/*Δ*t*) of the simulation, where Δ*t* is the sampling rate and *T* is the duration of the simulation, the velocity matrix is changed as:

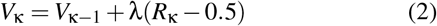

where λ can be referred as the momentum that controls the velocity change, and *R*_κ_ ∈ ℝ^*nx*2^ is a matrix of uniformly distributed random numbers in the range from 0 to 1. The velocity is zeroed for the particles at rest in quarantined, isolated, severely infected and dead states, or when it goes over *v*_*max*_. Normalization in the range of [-0.5, 0.5] is applied to the *R*_κ_ matrix to provide velocity updates with zero bias.

Using the velocity terms from (2), the position of the particles are updated at each iteration as:

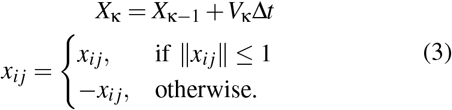

Particles are assigned an age group, *ag*, which enable differentiation of disease severity based on the age pyramid of the population of interest.

### B. Particle-based SEIR Simulator Model

As illustrated in Fig. 1, our particle-based SEIR simulator has four superstates: Susceptible (**S**^*s*^), Exposed (**E**^*s*^), Infected (**I**^*s*^) and Recovered (**R**^*s*^). The Exposed superstate (**E**^*s*^) consists of two states: Exposed (**E**) and Quarantined (**Q**), and the latter one has True (**TQ**) and False Quarantined (**FQ**) substates. The Infected superstate (**I**^*s*^) accommodates Infected (**I**), Isolated (**Iso**) and Severely Infected (**SI**) states, with the Isolated (**Iso**) state that has True (**TIso**) and False Isolated (**FIso**) sub-states. The last superstate, Recovered (**R**^*s*^) has the Vaccination Immunized (**VI**) and Recovery Immunized (**RI**) states. Vaccination Immunized state and the vaccination module is explained in detail in Section III-C. The list of parameters of the simulator are listed in Table II.

**Fig. 1:**
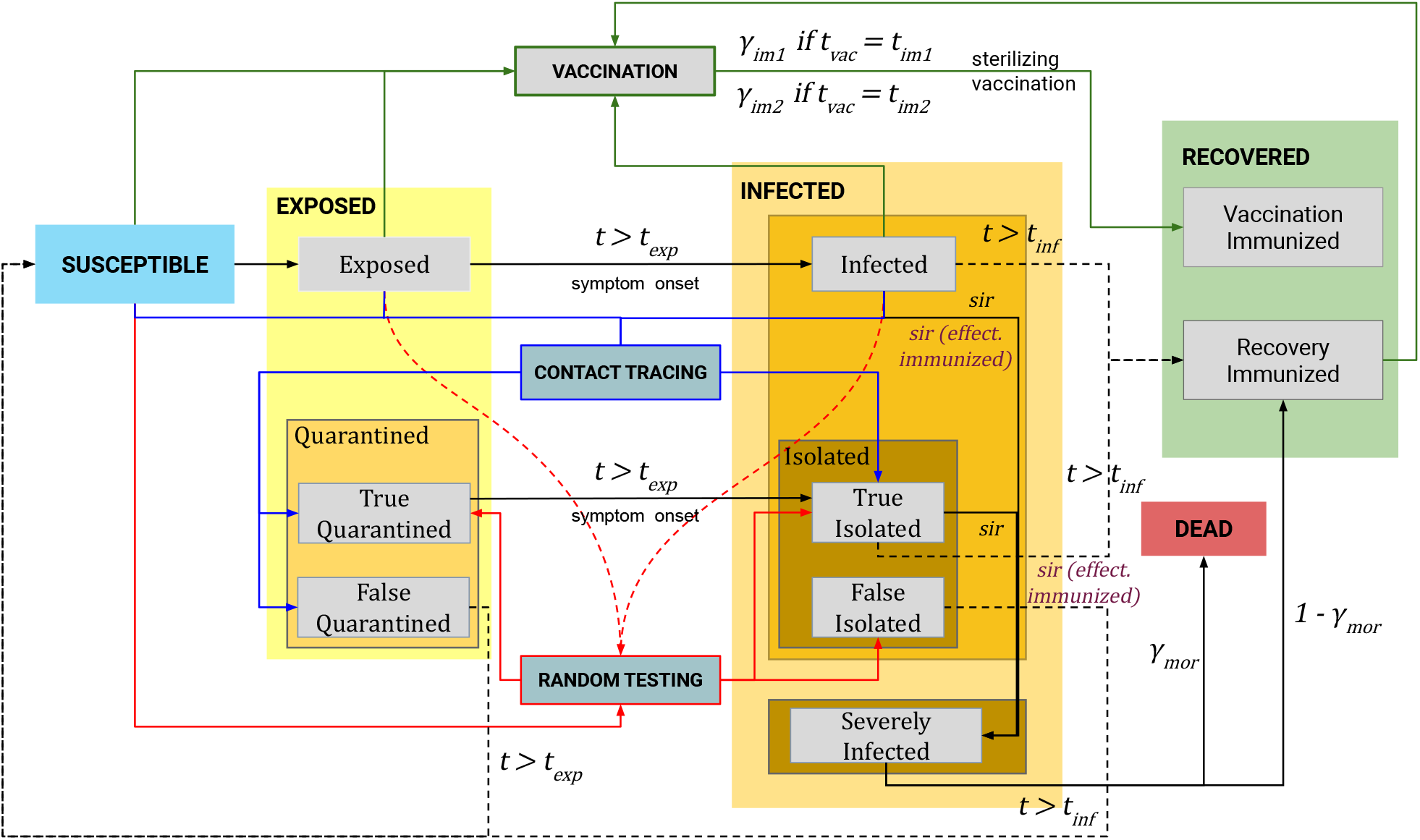
The state chart of the particle-based SEIR epidemic simulator with effective and sterilizing vaccination.

**TABLE II:** List of simulation parameters and their descriptions.

| Parameter | Description |
| --- | --- |
| $n$ | Total number of particles |
| $n_e$ | Initial number of exposed particles |
| $T$ | Simulation length in days |
| $x_{thr}$ | Minimum distance to transmit the disease |
| $v_{max}$ | Maximum allowed speed of particles |
| $\lambda$ | Speed gain |
| $sir$ | Daily rate of Infected/Isolated particles getting Severely Infected |
| $\gamma_{mor}$ | Severely Infected to Dead transition probability |
| $\epsilon_{exp}$ | Transmission probability of Exposed |
| $\epsilon_{qua}$ | Transmission probability of Quarantined |
| $\epsilon_{sev}$ | Transmission probability of Severely Infected |
| $t_{exp}$ | Exposure period in days |
| $t_{inf}$ | Infection period in days |
| $\beta$ | Ratio of the population using a contact tracing app |
| $\theta$ | Number of daily tests per thousand people |
| $sn$ | Sensitivity of tests |
| $sp$ | Specificity of tests |
| $\gamma_{im1}$ | First dose vaccination efficacy |
| $\gamma_{im2}$ | Second dose vaccination efficacy |
| $t_{vac}$ | Time of the first vaccination shot |
| $t_{im1}$ | First dose immunization period |
| $t_{im2}$ | Second dose immunization period |
| $\vartheta$ | Number of daily vaccines per thousand people |
| $sir_{ev}$ | Daily rate of Effective Immunized particles in the Infected/Isolated getting Severely Infected |

Initially, a certain number of particles *n*_*e*_ are chosen randomly and sent to the Exposed state, to seed the epidemic spread, and the rest are in the Susceptible (**S**^*s*^) superstate. Susceptible particles become exposed when they come in proximity to the particles spreading the infection (**E, TQ, I, SI** and **TIso**) closer than a predefined contact threshold *x*_*thr*_ distance; this proximity threshold is parameterized, and can be adjusted. The contagious particles spreading the infection vary in their transmission probability rates (ranging from 0 to 1) that are defined as ε_*exp*_ in case of exposed particles, ε_*qua*_ for for **TQ** and **TIso**, ε_*sev*_ for **SI**, respectively. After the period *t*_*exp*_, exposed particles move to the Infected state. In the Infected state, a certain proportion of particles transition to the Severely Infected state depending on the age-based *sir* parameter and the rest recover after the infection period *t*_*in f*_ is over. Among severely infected particles, some of them become deceased and others recover according to the mortality rate γ_*mor*_.

### C. Vaccination Modules

For our simulator, we developed a vaccination module with two post-vaccination immunization modes, to model sterilizing and effective immunity. As mentioned earlier, sterilizing immunity implies no further transmission of the virus to other individuals while the effective immunity obtained after vaccination reduces the probability of severe infection that might cause mortality. In either case, the vaccination can be modeled as either one shot or in two stages with a period in between, to account for the practice of two-dose COVID-19 vaccination. According to [11]–[13], the vaccine efficacy is determined by the number of doses administered and time after the vaccination. In our simulator, the vaccinated particles do not get additional doses after the second dose. In each of the modes, vaccination can be performed according to two strategies: random vaccination and age-based vaccination. In the random vaccination case, the age differences of the particles are not considered. In the age-based case the model takes into account particle age, in accordance with the stated strategy, e.g. the oldest particles are vaccinated first.

#### 1) Sterilizing Vaccination

In case of sterilizing vaccination, particles in the Exposed, Infected, Recovery Immunized, and Susceptible states are vaccinated according to the number of daily vaccines per thousand people ϑ. The vaccinated susceptible particles, that did not contact with the contagious particles, transition to the Vaccination Immunized state according to the first dose vaccination efficacy γ_*im*1_ when the immunization period *t*_*im*1_ is reached. Subsequently, when modeling the administration of the second-dose, the rest of the vaccinated susceptible particles transition to the Vaccination Immunized state based on the second dose vaccination efficacy γ_*im*2_ when the vaccination time reaches immunization time *t*_*im*2_. The remaining vaccinated particles do not gain immunity and continue with the regular flow of the simulation. The vaccination immunized particles are considered to achieve sterilizing immunity, thus, do not get infected and transmit the virus when they come in contact with the contagious particles. Susceptible particles, that were not immunized after the first dose, may get exposed before getting the second dose. In this case, they further transition according to the SEIR model states. Similarly, particles that did not transition to the Vaccination Immunized state after *t*_*im*2_ (the second dose) remain in the Susceptible state. For exposed, infected and recovered particles, the vaccination is considered to be wasted. Exposed and infected particles have the virus in their organism and, therefore, continue transitioning according to the SEIR states until recovery or death. As for recovery immunized particles, they remain in the Recovered state.

#### 2) Effective Vaccination

In the effective vaccination case, there is no Vaccination Immunized state in the Recovered superstate. Therefore, the susceptible particles immunized by the vaccine are designated as vaccination immunized, but they may still get infected with less probability of transitioning to Severely Infected sub-state (an asymptomatic or mild case of COVID-19). Similar to the sterilizing vaccination case, susceptible, exposed, infected and recovered particles get vaccinated. For the exposed, infected and recovered particles the vaccination is wasted, same as in the sterilizing vaccination. As for the susceptible particles, if they develop immunity either after the first or second dose (*t*_*im*1_ and *t*_*im*2_, respectively), their probability to get severely infected reduces to *sir*_*ev*_. If they don’t develop immunity and become exposed to the disease, they transition to the Severely Infected sub-state with *sir* rate. Effective immunized particles, which get infected and eventually recover, transition to the Recovery Immunized state.

### D. Simulator Implementation

We implemented our particle-based COVID-19 simulator with vaccination, contact tracing, and testing modules in MATLAB R2020. The source code is available on GitHub^1^ under MIT license. In addition, we provide the Python implementation of the simulator. All the simulations presented in the results section were performed in MATLAB.

## IV. Simulations and Results

### A. Validation Scenario for the Province of Lecco, Italy

In our previous work [4], we simulated the COVID-19 epidemic in the province of Lecco, Italy. We chose this region as it was an epicenter of the COVID-19 outbreak in Italy, for which comprehensive epidemic data is available, dating from February 24, 2020, shared by the Italian government [27].

In addition to the vaccination module, our enhanced simulator features age-based differentiation of the particles. Therefore, we first performed model calibration and validation of our new simulator based on the statistics for the province of Lecco. We set the number of particles to 337,087 according to the demographic statistics [26]. We started the simulation from January 1, 2020 with ten initial exposed particles. The model was configured for a 361 day period up to the start of vaccination in the province. The particles were randomly assigned to age groups according to the population pyramid presented in Table III. We set the age-based daily rate of infected/isolated particles getting severely infected, *sir*, equal to the age-based COVID-19 case fatality rates for Lombardy region as given in the Table III; we made this assumption as the province of Lecco is within the Lombardy region and the case fatality rates at the province level were not available. Age-based *sir* enabled us to differentiate the probability of a particle becoming severely infected, which results in higher mortality. As for the daily testing parameter θ, we used the daily tests per 1000 people based on the official statistics [28]. The β parameter that is used for contact tracing was set to zero.

**TABLE III:** Population pyramid and age-based epidemic fatality rate of province of Lecco, Italy.

| Age Group | Age Interval | Population | Population (%) | Case Fatality Rate (%) [26] |
| --- | --- | --- | --- | --- |
| 1 | 0-9 | 28,403 | 8.4 | 0 |
| 2 | 10-19 | 32,682 | 9.7 | 0.01 |
| 3 | 20-29 | 33,613 | 10.0 | 0.03 |
| 4 | 30-39 | 37,921 | 11.2 | 0.08 |
| 5 | 40-49 | 48,520 | 14.4 | 0.15 |
| 6 | 50-59 | 51,366 | 15.2 | 0.6 |
| 7 | 60-69 | 43,717 | 13.0 | 2.2 |
| 8 | 70-79 | 28,055 | 8.3 | 5.1 |
| 9 | 80+ | 32,811 | 9.7 | 9.35 |

In order to fine tune the parameters, we utilized the timeline of official governmental policies in the region (see Table IV). To imitate the lockdown, quarantine and lift up of certain restrictions, we decreased and increased the maximum allowed velocity of the particles *v*_*max*_ and momentum λ according the epidemic timeline. The rest of the calibrated parameters are summarized in Table V.

**TABLE IV:** Major events and NPIs in Lecco province during the COVID-19 epidemic [30]

| Day | Date | Event |
| --- | --- | --- |
| 0 | 1/1/2020 | Start of the simulation. |
| 55 | 24/2/2020 | The COVID-19 data repository was launched [27]. |
| 71 | 11/3/2020 | Lockdown in the province. Bars, restaurants are closed. |
| 82 | 22/3/2020 | Factories and all nonessential productions are closed. |
| 124 | 4/5/2020 | Easing lockdown between the regions. |
| 154 | 3/6/2020 | Unrestricted travel is allowed. |
| 162 | 11/6/2020 | International flights in Milan are resumed. |
| 197 | 15/7/2020 | International borders are closed. Restrictions are back. |
| 256 | 14/09/2020 | Schools are opened. |
| 290 | 17/10/2020 | Hybrid teaching in school. Food service restrictions. |
| 298 | 25/10/2020 | Nationwide restrictions. Night-time curfew [31]. |
| 326 | 23/11/2020 | Lombardy still in red zone. |
| 337 | 3/12/2020 | Classified as very high risk region. Strict regime until 15 January [32]. |
| 361 | 27/12/2020 | Start of vaccination. End of the simulation. |

**TABLE V:** Simulation Parameters for Lecco, Italy.

| $v_{max}$ (Days) | $\lambda$ (Days) | $x_{thr}$ (Days) | $sir$ (Age group) | $sp$ | $sn$ |
| --- | --- | --- | --- | --- | --- |
| 0.02 (0-55) |  |  | 0 (1) |  |  |
| 0.012 [55-71] |  |  | 0.01 (2) |  |  |
| 0.006 [71-82] |  |  | 0.03 (3) |  |  |
| 0.004 [82-154] | 0.002 (0-55) | 8.6e-5 (0-154) | 0.08 (4) | 0.95 | 0.99 |
| 0.02 [154-200] | 0.0012 [55-71] | 6.9e-5 [154-200] | 0.15 (5) |  |  |
| 0.007 [200-256] | 0.0006 [71-82] | 8.6e-5 [200-361] | 0.6 (6) |  |  |
| 0.012 [256-290] | 0.0004 [82-154] |  | 2.2 (7) |  |  |
| 0.01 [290-298] | 0.002 [154-365] |  | 5.1 (8) |  |  |
| 0.008 [298-337] |  |  | 9.35 (9) |  |  |
| 0.007 [337-361] |  |  |  |  |  |
| $t_{inf}$ | $t_{exp}$ | $\epsilon_{exp}$ | $\epsilon_{qua}$ | $\epsilon_{sev}$ | $\gamma_{mor}$ |
| 14 | 5 | 0.7 | 0.3 | 0.3 | 0.14 |

Figure 2 shows the calibration results for the province of Lecco, Italy. We observe the first epidemic peak from mid-March 2020 till the end of May 2020 followed by stabilization of the epidemic. We notice the epidemic surge starting from the end of September 2020, that was presumably triggered after the transition of education from online to hybrid format, which presumably increased social interaction amongst the students. The calibration simulation validates that our simulator can accurately replicate the effects of governmental policies on the epidemic dynamics of a certain region. As shown in the figure, the reported total of confirmed cases is around six times smaller than the simulation results, which is in agreement with the seroprevalence survey conducted by the local government [29]. The implementation dates of major NPIs in Table IV are indicated with dashed vertical lines along with the respective days for this calibration model. The total deaths according to the simulation and actual statistics for the period of interest are 797 and 830, respectively.

**Fig. 2:**
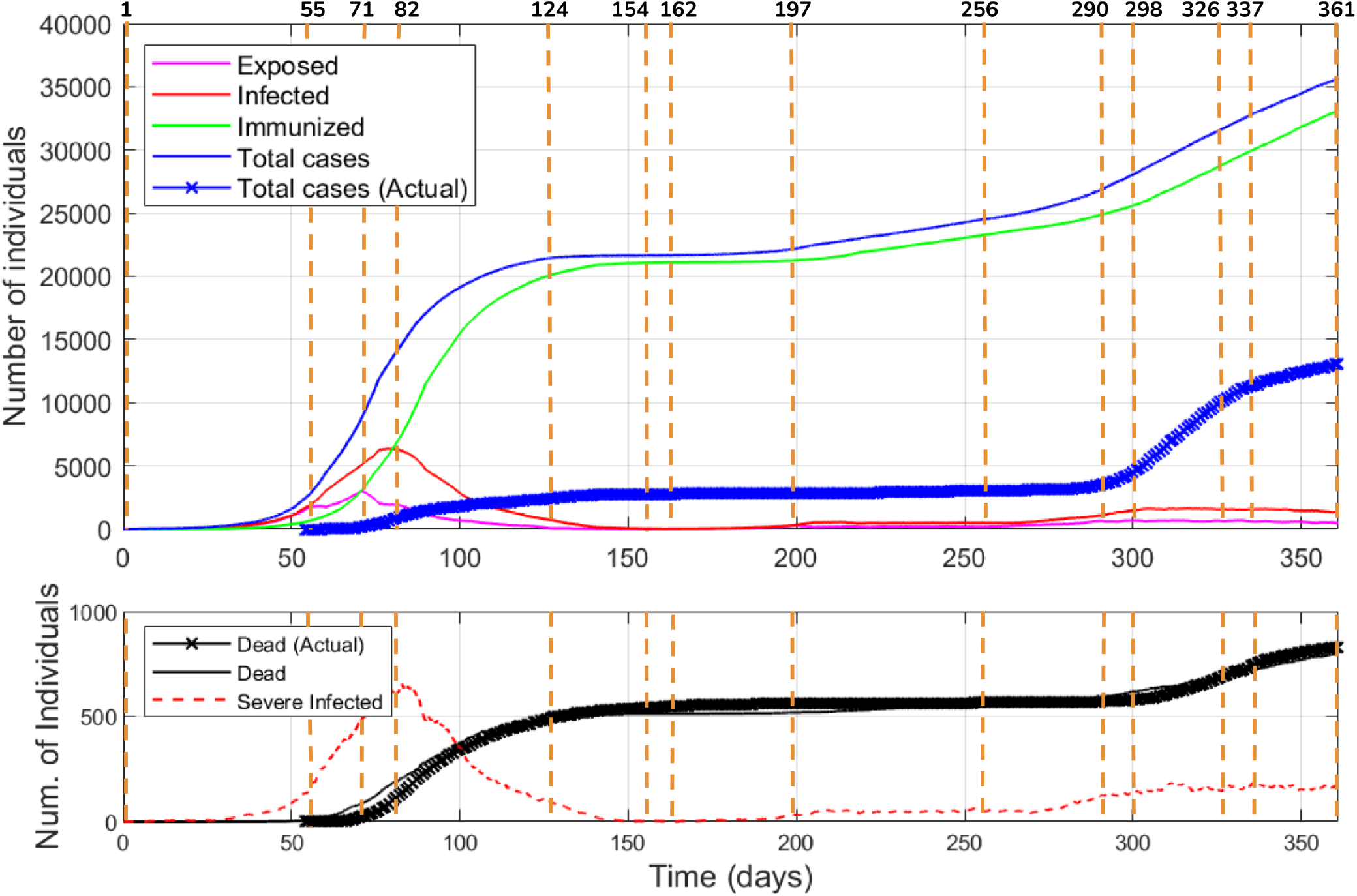
The results of the model calibration for the province of Lecco, Italy. The days of governmental policies are shown as vertical dashed lines and enumerated starting with the calibration start date (see Table IV). The upper plot shows epidemic states reported and simulated total cases versus time. The bottom plot shows the reported and simulation deaths as well as the dynamics of severely infected versus time

### B. Vaccination Strategies

We initialized the simulations for different vaccination strategies with the epidemic state reached in the end of the calibration simulation described in the Section IV-A.

We used the results of clinical studies of vaccines from Pfizer-BioNTech and Moderna as parameters for the vaccination module. The vaccines have age restriction of 18 and 16 year old, respectively. Since the age pyramid for the province of Lecco was available as per the age interval presented in Table III, we considered vaccination of age groups 3-9, i.e., 20 years old and above. Two doses are considered with a 21 day interval in between. The vaccine efficacy was set to γ_*im*1_ = 52% after *t*_*im*1_ = 12 days from the first vaccination date [11]. According to [11], [12], seven days after the second dose the vaccine efficacy reached 95%. We assume that particles that took the first dose will take the second dose too. Therefore, we used γ_*im*2_ = 95% at time *t*_*im*2_ = 28 days from the first vaccination date.

In subsections III-C1 and III-C2, we described the differences in the implementation of the sterilizing and effective vaccinations. In the sterilizing vaccination, the particles are sent to the Vaccination Immunized state based on the parameters of the vaccine given above. While in the effective vaccination, the immunization of particles implies a decrease in the daily rate of effective immunized particles in the Infected or Isolated states getting severely infected to *sir*_*ev*_. Since the vaccine efficacy in the real world requires time to observe, we assumed *sir*_*ev*_ to be 5% of the baseline *sir* listed in Table V.

To analyze the effects of different strategies, we performed 32 simulations which consist of the combinations of four different numbers of daily vaccines per thousand people (ϑ = 2, 4, 6, 8), four different cases (*effective-age-based* and *effective-all, sterilizing-age-based* and *sterilizing-all*), and two target populations (age groups 3-9 and 3-7). In case of the vaccination for target population for age groups 3-7, age groups 8 and 9 follow the normal SEIR transitions. To see the effect of vaccination, we performed additional base scenario without vaccination. To account for the stochastic nature of the simulator, for each scenario, we performed five simulations with the average of the results presented in Figs. 3, 4, 5, and 6. According to the official statistics, the daily vaccines per thousand people varies from zero to eleven by March 23, 2021. For example, the vaccination is being implemented at highest rates in Israel, United Arab Emirates, Chile, United Kingdom and United States (ϑ = 11, 8, 5, 5 and 4, respectively), whereas other countries have rates less than one [28]. Since the statistics is provided as average for the whole country, the vaccination rates might differ in regions. Therefore, we considered ϑ = 2, 4, 6, 8 in this paper.

**Fig. 3:**
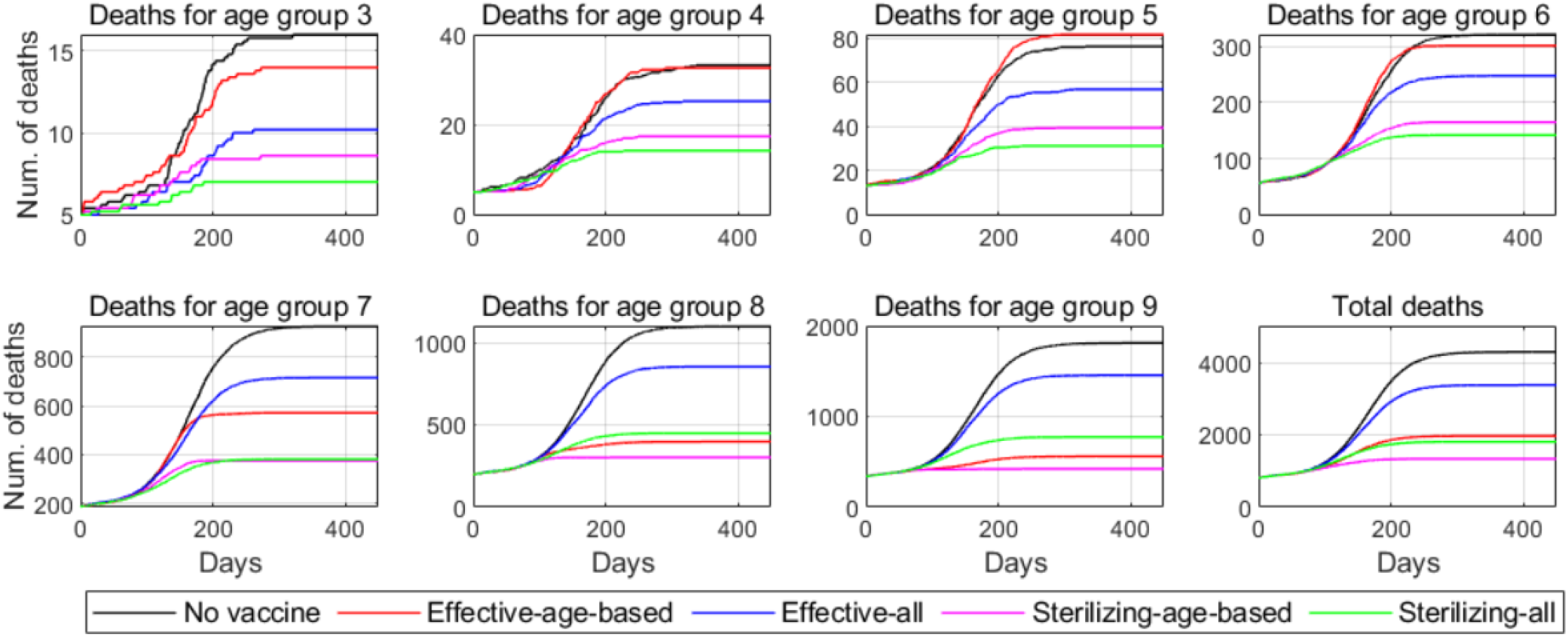
Comparative plots of four vaccination scenarios across the age group 3-9. The daily rate of vaccination per thousand people is 2. The target population for vaccination - age groups 3-9. Plots illustrate the transition of the best case across age groups: age-based vaccination is most optimal for groups 8 and 9. For age group 7 and below sterilizing-age-based and sterilizing-all scenarios result in similar numbers while the gap between the effective-age-based and effective-all cases increases with the ascending age group.

**Fig. 4:**
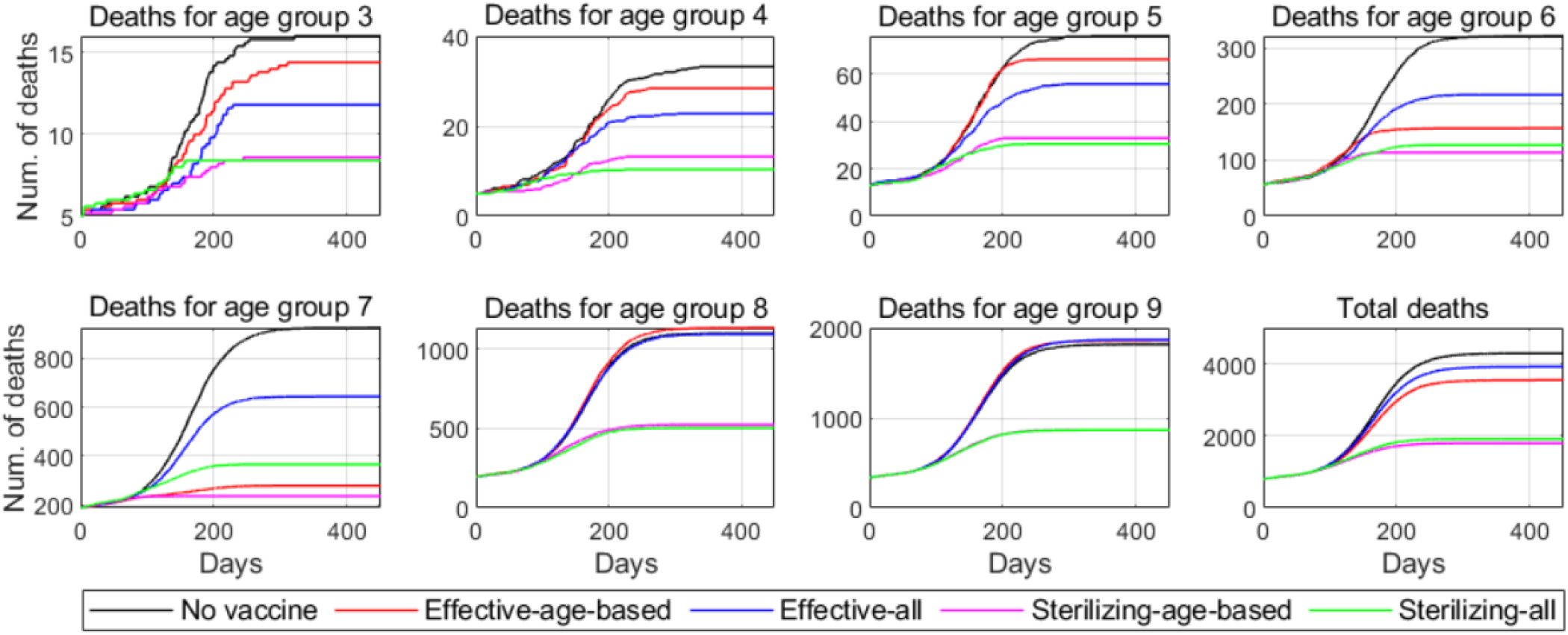
Comparative plots of four vaccination scenarios across the age groups 3-7. The daily rate of vaccination per thousand people is 2. The target population for vaccination is the age groups 3-7. The exclusion of the age groups 8 and 9 clearly illustrates a similar trend for groups 7 and below as in Fig. 3. Age groups 8 and 9 show that the sterilizing immunity of the rest of the populations has a substantial potential to achieve herd immunity and results in much smaller death numbers compared to the effective case.

**Fig. 5:**
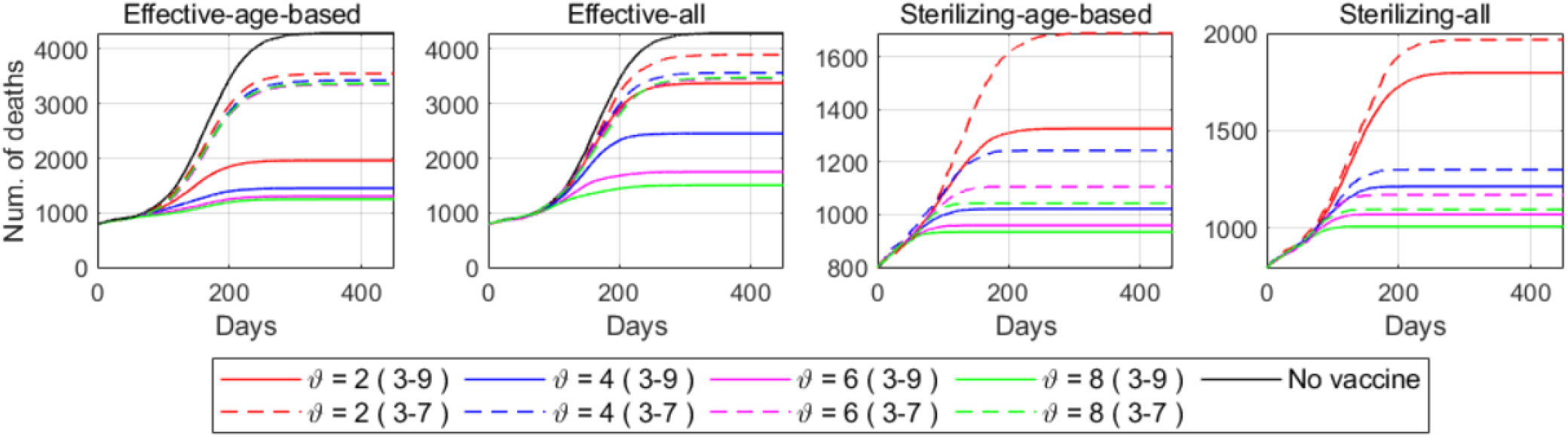
Comparative plots of deaths for different vaccination rates and target populations by four different scenarios: Effective-age-based, Effective-all, Sterilizing-age-based, and Sterilizing all. Effective vaccination cases clearly illustrate the effect of the restriction of the vaccination to certain age groups. The difference between the strategies is negligible in case age groups 8 and 9 are excluded from the vaccination. In the sterilizing scenario, the difference between the random-all and age-based vaccination decreases as the vaccination rate increases. The effect of the herd immunity can be seen in the sterilizing-all plots, where the numbers of deaths are much smaller compared to the effective-all cases.

**Fig. 6:**
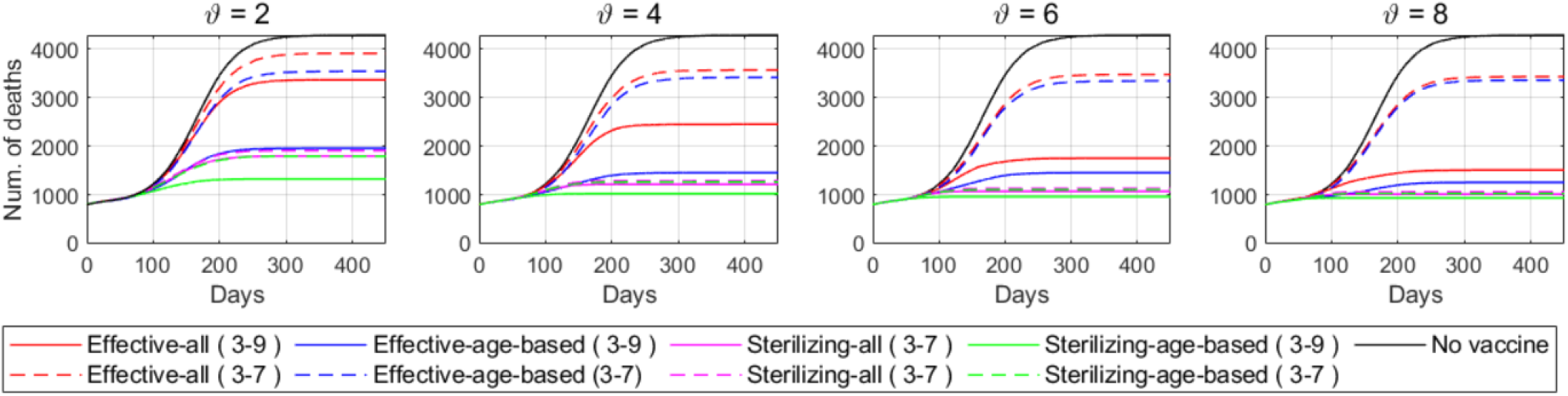
Comparative plots of age-based and random-all effective and sterilizing vaccination scenarios for different target populations by daily vaccination rates per thousand people = 2, 4, 6, and 8. For smaller vaccination rates the epidemic dynamics are highly sensitive to the vaccination scenario and immunization type. For the effective case, it is critical to vaccinate first the most vulnerable portion of the population.

We provide a video as a supplementary material which shows simulations for four scenarios with ϑ = 2: 1) *sterilizing-all*, 2) *sterilizing-age-based*, 3) *effective-all*, and 4) *effective-age-based*. These clips demonstrate the dynamics of the epidemic and the effects of different vaccination cases on a 2D map by age groups.

Figures 3 and 4 show the total deaths and deaths by age group for different strategies and target population at ϑ = 2. We observe that the *sterilizing age-based* vaccination provided lowest deaths followed by the *sterilizing-all*, 1,702 and 1,797, respectively. The *effective-age-based* vaccination also resulted in numbers close to the *sterilizing-all* (1,979 deaths). As we started vaccinating the most vulnerable group (from oldest) first which after immunization have considerably lower probability to get severely infected and deceased, the number of deaths was very small compared to the *effective-all* case with 3,371 deaths. The same trend is observed in Fig. 4, except in this case we vaccinated age groups 3-7, which illustrates less difference between no vaccine scenario, *effective-all* and *effective-age-based* as most of the deaths come from age groups 8-9. As we restrict the vaccination to age groups 3-7, the *sterilizing-all* and *sterilizing-age-based* resulted in similar numbers of deaths, 1,795 and 1,912, respectively.

The effectiveness of different strategies for each vaccination rate ϑ can be observed from Fig. 6. At lower ϑ, the difference between the effective and sterilizing vaccination cases is relatively large, but as the number of daily vaccines per thousand people increases the difference narrows.

The comparison of different vaccination rates for each strategy is illustrated on Fig. 5. For the sterilizing vaccination case, we did not include the scenario with no vaccine in the plot due to large difference (for better visualization). Effective vaccination shows a clear difference between the death numbers for two cases (age groups 3-7 and 3-9) compared to the sterilizing vaccination. A considerable difference is observed between the ϑ = 2 and ϑ = 4, respectively. The nonlinear relationship between the vaccination rate and decrease in mortality is noted. This gives an intuition to find an optimal number of daily vaccines per thousand people for certain population as a tradeoff between the vaccination supply, number of deaths, pressure on healthcare and definition of priority groups.

## V. Discussion

The COVID-19 virus and subsequent mitigation efforts have incurred significant social and economic damage, which could continue for some time to come. The initial response to the virus consisted primarily of NPIs, such as social distancing, masks, and varying degrees of social “lockdowns”. These measures were often successful, for short intervals, but when relaxed (or ignored) the epidemic resurged with exponential growth rates.

The emergence of vaccines has introduced a new critical challenge: the need for a customized strategy to achieve population immunization. It is not possible to vaccinate all people at once, given the bottlenecks of vaccine approvals, production, procurement, distribution, storage, dissemination, and monitoring. The situation is exacerbated by the exhausted state of the existing healthcare systems, and the need to balance competing objectives to achieve the desired goals.

Thus, it is necessary to identify the policy goals, and establish priority vaccination groups, in an effort to maximize the social and economic benefit, or at least, to reduce the worst-case outcomes.

The formation of vaccination priority groups also includes practical considerations such as the ability to acquire and administer the vaccinations, and whether there is a strong healthcare infrastructure, sufficient funding and a system for vaccination monitoring [8]. The common strategies are aimed to reduce the mortality, decrease the pressure on the healthcare, and minimize the infection spread over a specific period of time [5].

The vaccination rate is another critical parameter that determines the effectiveness of random or age-based vaccination strategies. According to several studies [5], [6], age-based vaccination is the most optimal solution to minimize the number of deaths in case of low vaccination rate which is consistent with the performed simulation results. Whereas for medium or fast vaccination rates, random vaccination might be preferable, as the younger generations can spread the infection widely until vaccinated and therefore result in an overload of the healthcare system.

If we examine each age group separately (see Fig. 3), we observe that for age groups 8-9 it is the *sterilizing-age-based* and *effective-age-based* cases that result in the fewest deaths, as in this case we prioritize vaccination of the high-risk elderly. As we descend down the age groups, however, we observe that the *sterilizing-all* and *sterilizing-age-based* cases produce the fewest death outcomes. Furthermore, across the age groups we see the change of order of different cases: the age-based vaccination strategies result in fewer deaths for older groups and random all strategy becomes more preferable as we descent down the age groups.

When vaccination is restricted to certain age groups, *sterilizing-age-based* and *sterilizing-all* results in very close deaths numbers for age groups excluded from the target vaccination (see Fig. 4). Similarly, *effective-age-based* yields the deaths number close to *effective-all* and no vaccination scenarios for excluded age-groups.

If we examine each case separately (see Fig. 5, we see that in *effective-age-based* and *effective-all* cases there is a clear offset between the strategies: vaccination of age groups 3-7 and 3-9. Whereas in the sterilizing vaccination case due to the condition that immunized particles do not further transmit the infection, we see that sterilizing immunity of the younger population helps to slow down the infection spread.

We see in Fig. 6 that as the vaccination rate increases, the difference in target population does not result in a big difference between the *sterilizing-all* and *sterilizing-age-based* strategies. Therefore, provided sufficient vaccine doses are available, the prioritization of certain population groups is not of critical importance and it is thus preferred to randomly vaccinate all population above the age of 19. But for effective immunization case, vaccinating the most vulnerable groups first is essential.

Table VI shows that, when we restrict the vaccination to age groups 3-7, the number of deaths is high since most of the deaths are from the oldest age groups 8-9, whom we are excluding from vaccination in this case. Furthermore, the increase in vaccination rate does not result in a dramatic decrease of the mortality (3,539 and 3,916 deaths at ϑ = 2 and 3,433 and 3,429 at ϑ = 8 for *effective-age-based* and *effective-all* cases). The vaccination and following immunization of the other particles do not result in considerable decrease in deaths because effective immunized particles are considered to be able to transmit the virus. Now, if we consider the results of the sterilizing case, the increase in the daily vaccines per thousand people results in a dramatic decrease of deaths for both age group cases 3-9 and 3-7 as immunized particles are no longer getting infected and transmit the infection (age groups 3-9: 1,702 and 1,797 at ϑ = 2 and 933 and 1,009 at ϑ = 8 for *sterilizing-age-based* and *sterilizing-all* cases, respectively; age groups 3-9: 1,795 and 1,913 at ϑ = 2 and 1,059 and 1,052 at ϑ = 8 for *sterilizing-age-based* and *sterilizing-all* cases, respectively).

**TABLE VI:** Deaths by age-group for 32 different scenarios for the province of Lecco, Italy.

| Daily vaccination rate for 1000 people $\vartheta = 2$, Age groups 3-9 | | | | | | | | |
| --- | --- | --- | --- | --- | --- | --- | --- | --- |
| Strategy | [20-29) | [30-39) | [40-49) | [50-59) | [60-69) | [70-79) | [80+) | Total |
| Effective-age-based | 14 | 33 | 82 | 302 | 571 | 397 | 557 | 1959 |
| Effective-all | 10 | 25 | 57 | 248 | 715 | 855 | 1458 | 3371 |
| Sterilizing-age-based | 9 | 17.4 | 39.4 | 165 | 381 | 300 | 418 | 1702 |
| Sterilizing-all | 7 | 14 | 31 | 142 | 381 | 448 | 771 | 1797 |
| Daily vaccination rate for 1000 people $\vartheta = 2$, Age groups 3-7 | | | | | | | | |
| Effective-age-based | 14 | 28 | 66 | 157 | 279 | 1134 | 1867 | 3539 |
| Effective-all | 12 | 23 | 56 | 217 | 644 | 1094 | 1867 | 3916 |
| Sterilizing-age-based | 9 | 13 | 33 | 114 | 236 | 524 | 1861 | 1795 |
| Sterilizing-all | 8 | 10 | 31 | 127 | 366 | 504 | 864 | 1913 |
| Daily vaccination rate for 1000 people $\vartheta = 4$, Age groups 3-9 | | | | | | | | |
| Effective-age-based | 13 | 29 | 60 | 164 | 318 | 323 | 541 | 1453 |
| Effective-all | 9 | 17 | 44 | 169 | 537 | 619 | 1051 | 2453 |
| Sterilizing-age-based | 7 | 10 | 21 | 94 | 247 | 253 | 388 | 1021 |
| Sterilizing-all | 6 | 7 | 23 | 87 | 274 | 301 | 513 | 1214 |
| Daily vaccination rate for 1000 people $\vartheta = 4$, Age groups 3-7 | | | | | | | | |
| Effective-age-based | 10 | 19 | 34 | 92 | 261 | 1106 | 1844 | 3418 |
| Effective-all | 8 | 12 | 35 | 138 | 420 | 1100 | 1854 | 3572 |
| Sterilizing-age-based | 6 | 7 | 22 | 76 | 214 | 348 | 573 | 1288 |
| Sterilizing-all | 7 | 8 | 20 | 137 | 257 | 347 | 574 | 1288 |
| Daily vaccination rate for 1000 people $\vartheta = 6$, Age groups 3-9 | | | | | | | | |
| Effective-age-based | 11 | 19 | 35 | 104 | 287 | 313 | 533 | 1307 |
| Effective-all | 8 | 11 | 31 | 119 | 382 | 438 | 760 | 1752 |
| Sterilizing-age-based | 7 | 7 | 21 | 78 | 232 | 232 | 378 | 959 |
| Sterilizing-all | 7 | 6 | 18 | 77 | 245 | 258 | 458 | 1071 |
| Daily vaccination rate for 1000 people $\vartheta = 6$, Age groups 3-7 | | | | | | | | |
| Effective-age-based | 8 | 12 | 24 | 90 | 267 | 1093 | 1845 | 3345 |
| Effective-all | 6 | 9 | 22 | 99 | 316 | 1100 | 1101 | 3476 |
| Sterilizing-age-based | 7 | 7 | 19 | 77 | 212 | 290 | 492 | 1297 |
| Sterilizing-all | 6 | 6 | 18 | 77 | 329 | 288 | 402 | 1128 |
| Daily vaccination rate for 1000 people $\vartheta = 8$, Age groups 3-9 | | | | | | | | |
| Effective-age-based | 9 | 12 | 25 | 95 | 273 | 307 | 526 | 1253 |
| Effective-all | 8 | 10 | 23 | 102 | 315 | 384 | 661 | 1506 |
| Sterilizing-age-based | 7 | 7 | 18 | 77 | 218 | 227 | 378 | 933 |
| Sterilizing-all | 6 | 6 | 17 | 73 | 232 | 246 | 426 | 1009 |
| Daily vaccination rate for 1000 people $\vartheta = 8$, Age groups 3-7 | | | | | | | | |
| Effective-age-based | 7 | 9 | 22 | 87 | 260 | 1101 | 1869 | 3433 |
| Effective-all | 6 | 9 | 24 | 95 | 294 | 1120 | 1878 | 3429 |
| Sterilizing-age-based | 5 | 9 | 16 | 69 | 207 | 1119 | 1877 | 1059 |
| Sterilizing-all | 5 | 6 | 16 | 74 | 226 | 267 | 463 | 1052 |

Based on the deaths obtained for the model of province of Lecco, we see a dramatic decrease in mortality between ϑ = 2 and ϑ = 4 (see Table VI). Further, as we increase the daily vaccines per thousand people the decrease in the number of deaths slows down. This may imply that considering the specific parameter used to make analysis for province of Lecco, Italy, the optimal vaccination rate in case of limited vaccine supply might lie somewhere between ϑ = 2 and ϑ = 4.

Based on the results, it is evident that simulation across a range of possible vaccination scenarios, incorporating local demographics and propagation trends, can be an important component in the development of an optimal vaccination strategy, on a national or regional basis.

## VI. Conclusions

We have developed an SEIR particle-based epidemic simulator to facilitate the analysis of effective and sterilizing vaccination. The particles are distinguished by age, which is a critical feature due to the demographic variability of population susceptibility and outcomes. Thus, the model can more accurately represent the impact of government policies and vaccination strategies based on the profiles of the population. In the development of epidemic suppression and mitigation strategies it is imperative to define the primary goals. For example, to determine whether the immediate objective is to reduce the mortality rate, or to slow the spread of infection, or to reduce the impact on the healthcare system. The identification of the goal(s) will help determine the most effective combination of methods, both non-pharmaceutical (lockdowns, masks, distancing, etc.) and pharmaceutical (vaccinations and testing).

As an illustrative case, we performed simulations for the Lecco province of Italy to demonstrate how the methods chosen can influence the outcomes. We calibrated the model based on the reported data, using the actual timeline of the governmental policies, and then applied both age-based and random-all vaccination simulations across a range of population groups with varying rates of vaccination.

According to the obtained results, at lower vaccination rates the epidemic outcome is very sensitive to the selected vaccination strategy and type of resulting immunization. As an example, it is possible to substantially reduce overall mortality using an age-based vaccination strategy, though by prioritizing older people it leads to an increase in rates of infection amongst younger (unvaccinated) people, thus illustrating the tradeoffs involved in achieving policy goals. Overall, the best performance was achieved in the scenario of age-based vaccination yielding sterilizing immunity; the outcome of real-world vaccination will likely be found somewhere on the continuum between the two extremes, effective and the sterilizing immunization.

The type of vaccination immunity in the case of COVID-19 is still unknown, thus, the conditions necessary to achieve herd immunity are difficult to reliably establish at this time. The complexities with vaccine supply, economic disparities across and within countries, and the wide variability in the adoption and effective observance of NPIs may delay the achievement of practical herd immunity, on both a global and localized basis. Under these circumstances, this vaccination simulation tool can provide an effective way to model and consider the outcome of various scenarios, based on the desired goals, with the ability to incorporate updated actual data in near-realtime to provide ongoing assessment of strategies and projected outcomes.

The simulator could be enhanced by including additional factors in the particle contact component, such as physical location and age differentiation of social interaction and its impact on the transmission rate attribute of a simulator particle. While these factors could probably increase the sensitivity of the simulator, it would make the parameterization of the simulator more challenging.

## Data Availability

N/A

https://www.youtube.com/watch?v=z2j4hcmmOwc&ab_channel=ISSAI_NU

https://github.com/IS2AI/Particle-Based-COVID19-Simulator

## Footnotes

1 github.com/IS2AI/Particle-Based-COVID19-Simulator

## References

[1] WHO. (2021) WHO Coronavirus Disease (COVID-19) Dashboard. Last accessed on 2021-03-13: https://covid19.who.int/.

[2] A. Kuzdeuov, D. Baimukashev, A. Karabay, B. Ibragimov, A. Mirzakhmetov, M. Nurpeiissov, M. Lewis, and H. A. Varol, “A network-based stochastic epidemic simulator: Controlling COVID-19 with region-specific policies,” IEEE Journal of Biomedical and Health Informatics, vol. 24, no. 10, pp. 2743–2754, 2020.

[3] T. S. Farthing and C. Lanzas, “Assessing the efficacy of interventions to control indoor SARS-Cov-2 transmission: An agent-based modeling approach,” medRxiv, 2021. [Online]. Available: https://www.medrxiv.org/content/early/2021/01/22/2021.01.21.21250240

[4] A. Kuzdeuov, A. Karabay, D. Baimukashev, B. Ibragimov, and H. A. Varol, “Particle-based COVID-19 simulator with contact tracing and testing,” IEEE Open Journal of Engineering in Medicine and Biology, vol. 2, pp. 111–117, 2021.

[5] B. Goldenbogen, S. O. Adler, O. Bodeit, J. A. Wodke, X. Escalera-Fanjul, A. Korman et al., “Optimality in COVID-19 vaccination strategies determined by heterogeneity in human-human interaction networks,” medRxiv, 2020. [Online]. Available: https://www.medrxiv.org/content/early/2020/12/18/2020.12.16.20248301

[6] C. R. MacIntyre, V. Costantino, and M. Trent, “Modelling of COVID-19 vaccination strategies and herd immunity, in scenarios of limited and full vaccine supply in NSW, Australia,” Vaccine, 2021.

[7] D. S. Kim, S. Rowland-Jones, and E. Gea-Mallorquí, “Will SARS-CoV-2 infection elicit long-lasting protective or sterilising immunity? Implications for vaccine strategies,” Frontiers in Immunology, vol. 11, p. 3190, Dec 2020, 33362759[pmid]. [Online]. Available: https://doi.org/10.3389/fimmu.2020.571481

[8] K. Hardt, P. Bonanni, S. King, J. I. Santos, M. El-Hodhod, G. D. Zimet et al., “Vaccine strategies: Optimising outcomes,” Vaccine, vol. 34, no. 52, pp. 6691–6699, 2016. [Online]. Available: http://www.sciencedirect.com/science/article/pii/S0264410X16310301

[9] R. Markovič, M. Šterk, M. Marhl, M. Perc, and M. Gosak, “Socio-demographic and health factors drive the epidemic progression and should guide vaccination strategies for best COVID-19 containment,” Results in Physics, vol. 26, pp. 104 433–104 433, Jul 2021, 34123716[pmid]. [Online]. Available: https://pubmed.ncbi.nlm.nih.gov/34123716

[10] I. Abdulrahman, “SimCOVID: Open-source simulation programs for the COVID-19 outbreak,” medRxiv, 2020. [Online]. Available: https://www.medrxiv.org/content/early/2020/06/22/2020.04.13.20063354

[11] F. P. Polack, S. J. Thomas, N. Kitchin, J. Absalon, A. Gurtman, S. Lockhart et al., “Safety and efficacy of the BNT162b2 mRNA COVID-19 vaccine,” New England Journal of Medicine, vol. 383, no. 27, pp. 2603–2615, 2020. [Online]. Available: https://doi.org/10.1056/NEJMoa2034577

[12] L. A. Jackson, E. J. Anderson, N. G. Rouphael, P. C. Roberts, M. Makhene, R. N. Coler et al., “An mRNA vaccine against SARS-CoV-2: Preliminary report,” New England Journal of Medicine, vol. 383, no. 20, pp. 1920–1931, 2020. [Online]. Available: https://doi.org/10.1056/NEJMoa2022483

[13] T. Burki, “The Russian vaccine for COVID-19,” The Lancet: Respiratory Medicine, vol. 8, no. 11, pp. e85–e86, 2020. [Online]. Available: https://www.thelancet.com/journals/lanres/article/PIIS2213-2600(20)30402-1/fulltext?ref=brianlovin.com

[14] H. E. Randolph and L. B. Barreiro, “Herd immunity: Understanding COVID-19,” Immunity, vol. 52, no. 5, pp. 737–741, 2020. [Online]. Available: http://www.sciencedirect.com/science/article/pii/S1074761320301709

[15] A. Fontanet and S. Cauchemez, “COVID-19 herd immunity: Where are we?” Nature Reviews Immunology, vol. 20, no. 10, pp. 583–584, Oct 2020. [Online]. Available: https://doi.org/10.1038/s41577-020-00451-5

[16] G. Forni, A. Mantovani, L. Moretta, R. Rappuoli, G. Rezza, and A. Bagnasco, “COVID-19 vaccines: Where we stand and challenges ahead,” Cell Death & Differentiation, vol. 28, no. 2, pp. 626–639, Feb 2021. [Online]. Available: https://doi.org/10.1038/s41418-020-00720-9

[17] S. M. Sawyer, “What can we expect from first-generation COVID-19 vaccines?” The Lancet, no. 10261, pp. 1467–1469, 2020. [Online]. Available: https://www.thelancet.com/article/S0140-6736(20)31976-0/fulltext

[18] M. Piraveenan, S. Sawleshwarkar, M. Walsh, I. Zablotska, S. Bhattacharyya, H. H. Farooqui et al., “Optimal governance and implementation of vaccination programmes to contain the COVID-19 pandemic,” Royal Society Open Science, vol. 8, no. 6, p. 210429, 2021. [Online]. Available: https://royalsocietypublishing.org/doi/abs/10.1098/rsos.210429

[19] V. Priesemann, R. Balling, M. M. Brinkmann, S. Ciesek, T. Czypionka et al., “An action plan for pan-European defence against new SARS-CoV-2 variants,” Lancet), vol. 397, no. 10273, pp. 469–470, Feb 2021, 33485462[pmid]. [Online]. Available: https://pubmed.ncbi.nlm.nih.gov/33485462

[20] M. O’Driscoll, G. Ribeiro Dos Santos, L. Wang, D. A. T. Cummings, A. S. Azman, J. Paireau et al., “Age-specific mortality and immunity patterns of SARS-CoV-2,” Nature, vol. 590, no. 7844, pp. 140–145, Feb 2021. [Online]. Available: https://doi.org/10.1038/s41586-020-2918-0

[21] S. M. Sherman, L. E. Smith, J. Sim, R. Amlôt, M. Cutts, H. Dasch et al., “COVID-19 vaccination intention in the UK: Results from the COVID-19 vaccination acceptability study (CoVAccS), a nationally representative cross-sectional survey,” Human Vaccines & Immunotherapeutics, pp. 1–10, 2020, pMID: 33242386. [Online]. Available: https://doi.org/10.1080/21645515.2020.1846397

[22] A. A. Malik, S. M. McFadden, J. Elharake, and S. B. Omer, “Determinants of COVID-19 vaccine acceptance in the US,” EClinicalMedicine, vol. 26, p. 100495, 2020. [Online]. Available: http://www.sciencedirect.com/science/article/pii/S258953702030239X

[23] A. A. Dror, N. Eisenbach, S. Taiber, N. G. Morozov, M. Mizrachi et al., “Vaccine hesitancy: The next challenge in the fight against COVID-19,” European Journal of Epidemiology, vol. 35, no. 8, pp. 775–779, Aug 2020. [Online]. Available: https://doi.org/10.1007/s10654-020-00671-y

[24] M. D. Patel, E. Rosenstrom, J. S. Ivy, M. E. Mayorga, P. Keskinocak et al., “Association of simulated COVID-19 vaccination and nonpharmaceutical interventions with infections, hospitalizations, and mortality,” JAMA Network Open, vol. 4, no. 6, p. e2110782, Jun 2021, 34061203[pmid]. [Online]. Available: https://pubmed.ncbi.nlm.nih.gov/34061203

[25] W. K. Wong, F. H. Juwono, and T. H. Chua, “SIR simulation of COVID-19 pandemic in Malaysia: Will the vaccination program be effective?” 2021.

[26] P. Buonanno, S. Galletta, and M. Puca, “Estimating the severity of COVID-19: Evidence from the Italian epicenter,” PLOS ONE, vol. 15, no. 10, pp. 1–13, 10 2020. [Online]. Available: https://doi.org/10.1371/journal.pone.0239569

[27] Presidenza del Consiglio dei Ministri - Dipartimento della Protezione Civile. (2021) Dati COVID-19 Italia. Last accessed on 2021-02-10: https://github.com/pcm-dpc/COVID-19.

[28] E. Mathieu, H. Ritchie, E. Ortiz-Ospina, M. Roser, and J. Hasell, “A global database of COVID-19 vaccinations,” Nature Human Behaviour, vol. 5, no. 7, pp. 947–953, Jul 2021. [Online]. Available: https://doi.org/10.1038/s41562-021-01122-8

[29] Ministero della Salute. (2020) Primi risultati dell’indagine di siero-prevalenza SARS-CoV-2. Last accessed on 2020-10-2: http://www.salute.gov.it/imgs/C_17_notizie_4998_0_file.pdf.

[30] Garda World 2021. Italy authorities tighten COVID-19 measures from November 5. Last accessed on 2021-02-10: https://www.garda.com/crisis24/news-alerts/396716/italy-authorities-tighten-covid-19-measures-from-november-5-update-48.

[31] Medical Express. Italy’s Lombardy, Campania prepare virus curfews as cases jump. Last accessed on 2021-02-10: https://medicalxpress.com/news/2020-10-italy-lombardy-region-impose-virus.html.

[32] Law Business Research. Italy: COVID-19 - New restrictive measures enacted - Decree of December 3, 2020. Last accessed on 2021-02-10: https://www.lexology.com/library/detail.aspx?g=9a807adc-2d04-4e4c-b575-70a38d828666.

